# Artificial Intelligence in Early Detection of Autism Spectrum Disorder for Preschool ages: A Systematic Literature Review

**DOI:** 10.1101/2025.09.10.25335333

**Authors:** Hanan H. Hasan

## Abstract

**Background:** Early detection of autism spectrum disorder (ASD) improves outcomes, yet clinical assessment is time-intensive. Artificial intelligence (AI) may support screening in preschool children by analysing behavioural, neurophysiological, imaging, and biomarker data.

**Aim:** To synthesise studies that applied AI in ASD assessment and evaluate whether the underlying data and AI approaches can distinguish ASD characteristics in early childhood.

**Methods:** A systematic search of 15 databases was conducted on 30 November 2024 using predefined terms. Inclusion criteria were empirical studies applying AI to ASD detection in children aged 0–7 years. Reporting followed PRISMA 2020.

**Results:** Twelve studies met criteria. Reported performance (AUC) ranged from 0.65 to 0.997. Modalities included behavioural (eye-tracking, home videos), motor (tablet/reaching), EEG, diffusion MRI, and blood/epigenetic biomarkers. The largest archival dataset (M-CHAT-R) achieved near-perfect AUC with neural networks. Common limitations were small samples, male-skewed cohorts, and limited external validation.

**Conclusions:** AI can aid early ASD screening in infants and preschoolers, but larger and more diverse datasets, rigorous external validation, and multimodal integration are needed before clinical deployment.

## 1. Introduction

Autism Spectrum Disorder (ASD) is a neurodevelopmental condition that affects communication, behavior, and social interaction (APA, 2022). ASD affects an estimated 1 in 36 8-year-olds in the US, with substantial geographic variation, and about 1 in 100 children globally. Prevalence remains markedly higher in AMAB than AFAB children (male–female ratio ≈3.4–3.8 in recent US surveillance), with converging evidence that girls are under-recognized or diagnosed later, partly due to phenotype differences and masking. Early detection is critical for improving developmental outcomes, yet current diagnostic methods rely heavily on subjective behavioral evaluations which are time-intensive and require trained professionals (Kosmicki et al., 2015). Furthermore, the heterogeneity of ASD symptoms and the lack of reliable biomarkers complicate diagnosis, often delaying intervention (Stainbrook et al., 2019). These challenges are particularly pronounced in underserved areas with limited access to specialists.

Artificial intelligence (AI) has emerged as a transformative tool in healthcare, with successes in fields such as radiology, where AI models achieve diagnostic accuracies exceeding 90% (Rauschecker et al., 2020). In ASD diagnostics, AI techniques such as machine learning and deep learning offer the potential to analyze complex data types, including behavioral patterns, neuroimaging, and biomarkers, facilitating earlier and more accurate detection. For instance, machine learning models analyzing eye-tracking data have achieved diagnostic accuracies exceeding 84%, demonstrating AI’s potential in ASD detection (Wei et al., 2024). However, integrating AI into clinical workflows raises critical challenges, including data privacy, biases in AI models, and scalability (Natraj et al., 2024). Given the vulnerability of ASD populations and the sensitive nature of behavioral and neuroimaging data, addressing these ethical issues is essential for equitable adoption.

Despite the growing body of research on AI in ASD diagnostics, there is limited synthesis of its applications across multiple data modalities, such as combining neuroimaging with behavioral data for improved accuracy. Early detection is particularly critical during the infancy to preschool age range, a developmental period where interventions have the greatest potential to improve long-term outcomes (Lipkin et al., 2020).

This systematic review specifically focuses on this age group to assess how AI-driven tools have been applied to detect subtle markers of ASD, such as gaze anomalies, motor irregularities, and neurophysiological patterns. Understanding the extent to which AI can support early detection during this critical window is essential for advancing diagnostic methodologies. This systematic review aims to bridge this gap by critically evaluating AI-based approaches for early ASD detection, highlighting their strengths, limitations, and potential for clinical integration.

## 2. Methodology

The search engine EBSCOhost is used, where databases (APA PsycInfo, APA PsycTests, British Education Index, Business Source Premier, Child Development & Adolescent Studies, CINAHL Plus with Full Text, Education Abstracts (H.W. Wilson) Educational Administration Abstracts, ERIC, GreenFILE, Library, Information Science & Technology Abstracts, MEDLINE, Professional Development Collection, Psychology and Behavioral Sciences Collection, SPORTDiscus with Full Text, Education Research Complete) searched.

Search terms were combinations of key concepts related to AI, ASD, and early screening. Boolean operators and wildcard symbols were employed to maximize coverage and capture variations in terminology. Keywords (“artificial intelligence or ai or a.i. or machine learning or deep learning^*^” AND “early screening or early detection or early diagnosis or early identification” AND “autis^*^ or asd or autism spectrum disorder”) were searched.

A multi-phase selection process was conducted following PRISMA guidelines. Initial searches yielded 429 studies. Excluding the duplicates, 306 returned. The type of research has to be primary literature. The return of this default was 52 papers. Then in the title and abstract screening, any title that is not relevant to the topic is removed and the abstract which did not include any outcomes of detecting ASD or using other modalities than AI were excluded. A final selection of 12 studies was included in the analysis based on relevance, methodological rigor, and adherence to inclusion criteria. The PRISMA flowchart detailing these steps is referenced and visually represented for transparency in the figure (Fig. 1).

**Fig. 1.**
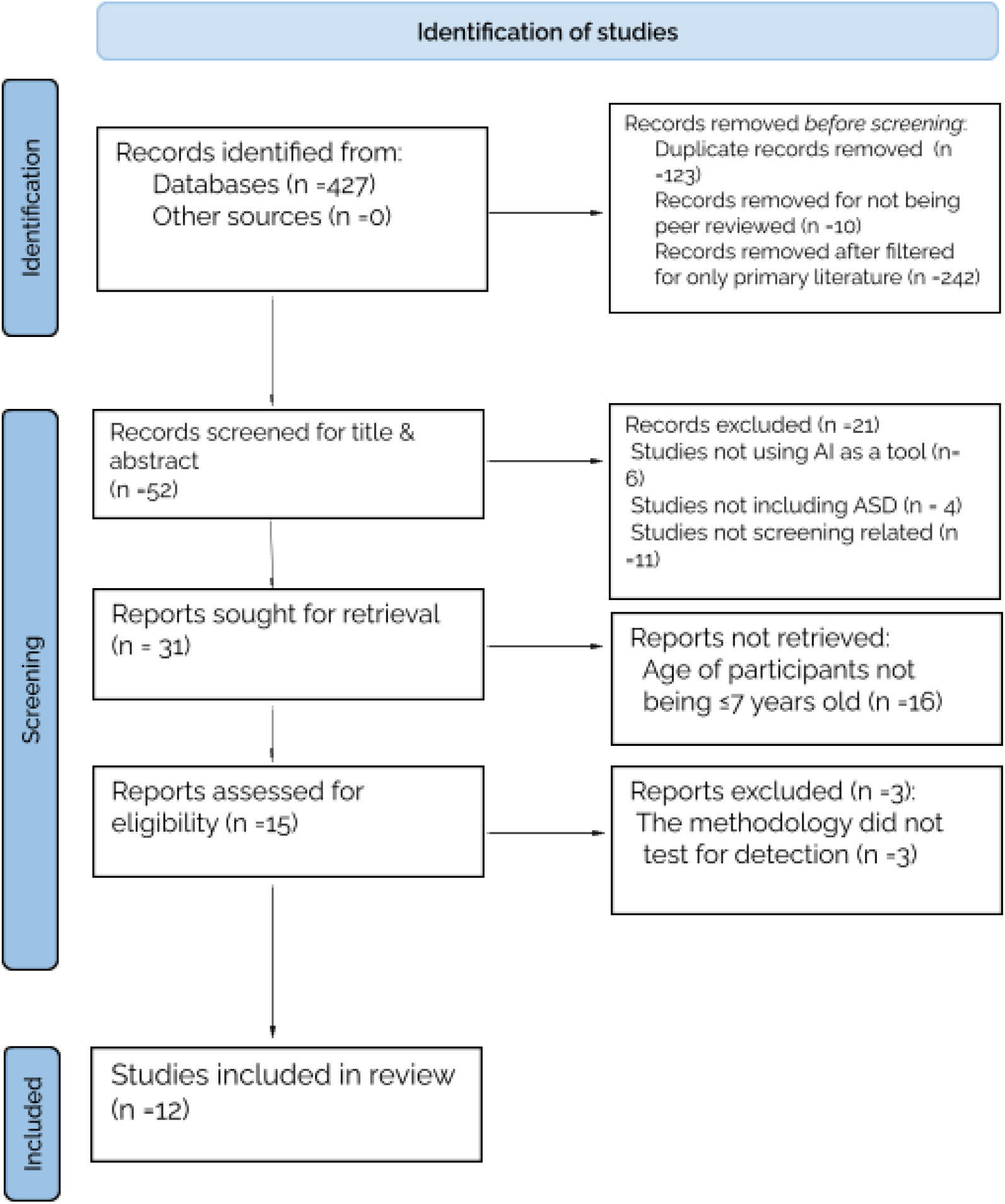
PRISMA 2020 flow diagram

### 2.1. Inclusion and Exclusion Criteria

This review included peer-reviewed studies published in any language or region. Non-English studies were included to ensure inclusivity, although all final included had an English version. The search was done on 30th November 2024, and the date range was unrestricted. search returned articles ranging from 2004 to 2024, reflecting the relatively recent development of AI applications in ASD diagnostics. Age specifications were put forth *from* birth to preschool. Limiting the scope to this age group allowed for an assessment of how AI-driven tools have been applied to enhance early detection during this pivotal stage. Studies were eligible if they focused on the application of AI in diagnosing ASD, particularly in children, and included empirical evidence of methods or tools.

Exclusion criteria encompassed studies doing logistical uses of AI without diagnostic focus, those lacking empirical data, or reviews and meta-analyses without original contributions. This strict inclusion strategy ensured that only high-quality, relevant studies were analyzed.

No formal tool was applied; studies were assessed on sample size, transparency, and data quality. Future work should adopt standardized tools (e.g., PROBAST/CASP)

### 2.2. Data Analysis

Key data were extracted from each study, including publication year, sample sizes, methodologies, AI techniques employed, and diagnostic metrics. The primary diagnostic metric, the Area Under the Curve (AUC), was used to evaluate performance, with values ranging from 0.65 to 0.997 across included studies. Sensitivity, specificity, and scalability were also considered. Although a formal quality appraisal framework was not used, studies were assessed for reliability and relevance based on sample size, methodological transparency, and data quality. Future reviews should adopt standardized appraisal tools to enhance methodological rigor and bias assessment. The above information is summarized as shown in the following table (Summary Table).

## 3. Results

The application of AI in early ASD detection has demonstrated significant promise, with studies leveraging diverse methodologies, input data types, and evaluation metrics. The reviewed studies collectively highlight the potential of AI for early ASD detection, but they also reveal divergences in methodologies, population demographics, and performance across biomarkers, behavioral data, neuroimaging, neurophysiological data, and motion trajectories which are the inputs that are assessed in the studies.

This section compares, critically evaluates and integrates the findings, highlighting trends, gaps, and potential areas revealed in this topic by the literature.

### 3.1. Populations and Study Designs

The studies reviewed encompassed diverse population sizes, with a total of 112,158 children of ages birth–7, from small case-control cohorts (e.g., 26 participants in motion studies by (Luongo et al., 2024)) to large datasets like the 14,995 records in (Achenie et al., 2019). Gender ratios, where reported, skewed heavily male, reflecting ASD’s higher prevalence in diagnosis in males (Barone et al., 2018);(Wei et al., 2024). Age distribution focused predominantly on newborns, preschoolers, and young children, with 2 studies including school children (Emanuele et al., 2021), reflecting the emphasis on early detection and ensuring the importance of training AI models of such an age period for early screening of ASD.

Furthermore, diversity of study types is obvious among the studies, ranging from observational and experimental with control groups, cross-sectional, to retrospective cohorts, seen in (Summary Table 1). A notable advantage of cohort and experimental studies is their ability to address age-related concerns in early screening for children by focusing on the children’s eventual diagnosis of ASD. Small sample sizes in studies like (Luongo et al., 2024) raise concerns about overfitting, while larger datasets (Rahman et al., 2020) offer statistical power but lack behavioral insights.

**Table. 1.** Summary Table 2020 flow diagram

| Author | Year | Study Type | Sample Size (ASD/Control) | Age Group | Input Data | Methodology | (AUC) | AI Method | Key Findings |
| --- | --- | --- | --- | --- | --- | --- | --- | --- | --- |
| Barone et al. | 2018 | Case-Control | 42/35 | ≤5 years | Blood and Urine collection of 45 Metabolites | Dried blood spots analyzed via high-throughput methods. Naïve Bayes classifier identified significant acyl-carnitines. | 0.73 | Naïve Bayes | Identified metabolic profiles (low vitamin D, high citrulline levels, features of primary mitochondrial disease) for ASD. |
| Achenie et al. | 2019 | Retrospective | 14,995 (archival) | 16–30 months | M-CHAT-R Data, follow up questionnaire | Feedforward neural network (FNN) trained on archival M-CHAT-R responses, outperforming manual scoring methods. | 0.997 | FNN | Machine learning can be used as an accurate and potentially improved method of M-CHAT-R analysis and M-CHAT-R Follow-Up. |
| Bahado-Singh et al. | 2019 | Case-Control | 14/10 | Newborns | Genomic DNA from neonatal dried blood | Genome-wide methylation data analyzed with a neural network (NN) for classification. | 0.975 | NN | Methylation biomarkers of loci on genes of leukocytes of newborns subsequently diagnosed for ASD. |
| Wei et al. | 2024 | Retrospective | 529 | 3–6 years | Via binocular eye tracker tested smooth pursuits, visual search, and joint attention. Reflected social cognition of the child. | Retrospective eye-tracking dataset analyzed with Random Forest for gaze patterns across social and non-social stimuli. | 0.849 | Random Forest | Non-social gaze features (decreased pursuit duration towards non-social stimuli) is critical. Machine learning with eye-tracking data assists through ET assessment to identify children with ASD before the age of three in 3 minutes. |
| Paolucci et al. | 2023 | Observational | 10/22 | 9 - 18 months | Home Videos by parents or caregivers when the children were aged 9–18 months. | ML Analyzed social interactions in home videos using SHAP values for key interaction characteristics. | 0.914–0.938 | ML + SHAP | Interaction patterns identified ASD. |
| Luongo et al. | 2024 | Case-Control | 10/10 | 3–6 years | Tablet-based game of drag and drop task | Game-based tablet task captured motion trajectories. Artificial NN trained on acceleration and velocity features. | 0.93 | ANN | Acceleration crucial for detection. |
| Jin et al. | 2015 | Cohort | 40/40 | 6–24 months | Diffusion MRI | Diffusion MRI evaluated white matter connectivity patterns. Multi-kernel SVM classified ASD infants. | 0.8 | Multi-Kernel SVM | Detected early white matter changes. Discriminative connections identified which may serve as potential imaging connectomic biomarkers for ASD diagnosis or prediction. |
| Rahman et al. | 2020 | Case-Control Cohort | 1,397/94,741 | Child (birth–2015), Parents of children born 1997–2007 until 2015 | Electronic Medical Records (EMR) Data of child and parents | EMR analyzed with logistic regression to predict ASD risk factors based on parental and child demographic features. | 0.709 | Logistic Regression, artificial neural networks, random forest | Risk modeling (sociodemographic risk factors, medication groups, nutritive supplements, reproductive hormones) with parental data, combinations of features do appear predictive of ASD. |
| Emanuele et al. | 2021 | Case-Control | 13/13 | 3.6–7 years | Reaching-Grasping Motion recorded by infrared camera | Motion patterns evaluated using Support Vector Machine (SVM). Tasks included reaching-grasping movements. | 0.947 | SVM | Motor-based early recognition is feasible and effective in ASD. |
| Natraj et al. | 2024 | Case-Control Longitudinal | 80/80 | 1–5 years | ADOS Videos & ADOS Audio | Neural networks (NN) independently analyzed video and audio streams. Ensemble methods used for classification. | 0.789 | Video-Audio Multimodal NN | Multimodal features improve detection. NN is effective in mentioning different features that are currently under the standard ASD assessment. |
| Peng et al. | 2021 | Experimental | 32/32 | 3–6 years | EEG features | EEG signals under affection-evoked movie clips were analysed for positive and negative emotions with SVM, incorporating L1-norm feature selection for optimization. | 0.938 (positive emotion), 0.875 (negative emotion) | SVM + L1-Norm | EEG Feature Selection Algorithm show accuracy for emotion-based screening of children with ASD |
| Ke et al. | 2024 | Retrospective Cross-sectional | 246/42 | 2–4 years | EEG features (power, connectivity) | Machine learning algorithms trained on EEG biomarkers (5 features included) | 0.65 | Logistic Regression | Elevated gamma and beta coherence identified as ASD-specific features; moderate accuracy achieved. |

The under representation of females, despite known gender differences in ASD presentation, introduces a significant bias in model training and application. This gender imbalance could lead to lower diagnostic accuracy for females, who often present with subtler or atypical ASD traits. Furthermore, the lack of ethnic diversity in datasets raises concerns about the applicability of findings to non-Caucasian populations, potentially perpetuating health inequities. Addressing these biases requires deliberate efforts to recruit diverse cohorts in future research.

### 3.2. Data Sources and Relevance

Use of AI in mental health is a new topic just like the new rise of it. The studies are looking for a diverse array of matter with diverse features and tools. Among the features looked at are the demographic aspects of ASD, standard definitions of ASD like behavioural data, motor, available screening/diagnosing tools of neuroimaging/signaling, and biomarkers that may enhance earlier screening (genetic and blood biomarkers). All of which even need further in depth researching, enriching the literature for possible promising power they may serve.

#### 3.2.1. Biomarkers

Biomarkers have shown strong predictive capabilities in ASD diagnostics.(Bahado-Singh et al., 2019) utilized DNA methylation patterns in newborn leucocytes, achieving a high AUC of 0.975 with a neural network model. The study underscored the role of epigenetic markers in ASD-related neural development, highlighting their strong predictive potential. (Barone et al., 2018) focused on blood metabolites and achieved a moderate AUC of 0.73 with a Naïve Bayes model, identifying metabolic markers linked to oxidative stress and mitochondrial dysfunction, both associated with ASD.

Despite their promise, these studies share key limitations. Both relied on demographically homogeneous samples, lacking gender-specific and ethnically diverse analyses, which raises concerns about the generalizability of their findings. Additionally, biomarkers require invasive sample collection and specialized laboratory infrastructure, limiting their feasibility for widespread clinical use. Integrating biomarkers with less invasive data sources, such as behavioral or motor assessments, offers a practical approach to overcoming these challenges. Multimodal models could retain the precision of biomarkers while improving accessibility.

#### 3.2.2. Behavioral Data

Behavioral datasets, including eye-tracking and home video analyses, offered non-invasive and accessible options for ASD screening. Both eye-tracking and video analysis emphasize interaction patterns as key markers but differ in contextual breadth and scalability.

Eye-tracking studies (Wei et al., 2024) identified non-social gaze patterns as critical markers of ASD using binocular eye trackers. Wei et al. (2024) reported significant differences in gaze fixation and scanning between ASD and typically developing (TD) children, achieving an AUC of 0.849 using a Random Forest classifier analyzing responses to social and non-social stimuli. Home video analysis, leveraging ML techniques, provided high accuracy (Paolucci et al., 2023). Paolucci et al. utilized explainable AI on home video data recorded by parents or caregivers when the children were aged 9–18 months looking for interaction patterns identifying ASD. They report a higher AUC of 0.938. These differences may reflect the broader context captured in video data versus the controlled settings of eye-tracking. While video analysis achieved higher accuracy, involves privacy concerns and extensive preprocessing requirements, which may limit its scalability compared to eye-tracking methods.

Additionally, behavioral approaches are often influenced by environmental and cultural variability. For instance, differences in social norms and caregiver-child interaction patterns may impact the consistency of behavioral markers.

#### 3.2.3. Motor Data

Motion data from tablet-based tasks and game-based assessments has demonstrated strong potential in detecting atypical motor patterns in children with ASD. (Luongo et al., 2024) utilized a game-based tablet task to capture motion trajectories, training Artificial Neural Networks (ANNs– type of DL) on features like acceleration and velocity, achieving an AUC of 0.93. The study emphasized acceleration as a critical marker for ASD detection. Similarly, (Emanuele et al., 2021) analyzed reaching-grasping tasks using Support Vector Machines (SVMs– a ML model), achieving an AUC of 0.947. Both studies consistently identified key motion markers, such as reduced inter-joint coordination and impaired synergies, as significant indicators of ASD.

Features like reduced inter-joint coordination and impaired synergies are particularly significant, as they reflect motor delays and impairments commonly associated with ASD. However, findings from these studies are limited by small sample sizes (n=20 and n=26, respectively) increasing risks of overfitting. While the lack of demographic diversity limits the applicability of findings across broader populations. Despite these constraints, the high AUC scores highlight the potential of motion data to serve as a reliable marker for ASD detection, offering valuable insights into the motor irregularities associated with the disorder.

Motion data complements other approaches, such as behavioral and neuroimaging data, by providing non-invasive and objective insights into motor irregularities, which are critical for early ASD screening.

#### 3.2.4. Neurophysiological Signals and Neuroimaging Data

Neuroimaging and neurophysiological studies highlighted ASD-specific brain activity patterns but differed in their resource intensity. EEG studies offered promising insights into ASD-specific emotional and cognitive responses (Ke et al., 2024; Peng et al., 2021). Both studies showed that ASD children have unusual patterns of brain activity, especially in gamma frequency bands. EEG-based approaches demonstrated screening AUC of up to 0.937 under positive emotional stimuli, revealing atypical brain activity patterns in gamma frequency bands, a hallmark of ASD-related neural irregularities. However, Ke et al. looked at resting brain activity, while Peng et al. studied how the brain reacts to emotions.

Despite these promising findings, the reliance on controlled experimental setups and limited sample sizes restricts the scalability of EEG-based models from them.

Similarly, the diffusion MRI (dMRI) study of (Jin et al., 2015) demonstrated potential for identifying neural biomarkers, with

Multi-Kernel SVM models achieving an AUC of 0.76. Jin et al. (2015) focused on structural neural markers, leveraging complex white matter (WM) connectivity networks to provide detailed brain connectivity maps. These structural insights complement EEG’s dynamic functional analyses but may lack the spatial resolution offered by dMRI. The multiscale approach is a unique strength, although challenges like high computational demands and small sample sizes are notable limitations.

#### 3.2.5. Existing ASD Diagnostic Tools

The reviewed studies highlight AI’s integration into established ASD diagnostic tools, enhancing their precision and scalability. (Achenie et al., 2019) improved the M-CHAT-R questionnaire’s scoring with AI, achieving an AUC of 0.997, significantly reducing human error and streamlining the screening process. (Rahman et al., 2020) used machine learning with electronic medical records (EMR) to predict ASD risk factors, achieving an AUC of 0.709. This method leveraged routinely collected demographic and medical history data but faced challenges due to variability in data quality across EMR systems. (Natraj et al., 2024) applied multimodal neural networks to Autism Diagnostic Observation Schedule (ADOS) data, incorporating video and audio analyses to achieve an AUC of 0.789. This innovative approach combined traditional assessments with automated neural networks to capture nuanced behavioral and communicative features. However, these studies also revealed limitations. The reliance on high-quality, standardized data sources, such as annotated ADOS recordings or EMR, limits across diverse populations and healthcare settings. Moreover, integrating AI into these frameworks requires addressing data biases and ensuring accessibility to advanced technological resources.

Despite these challenges, these advancements underscore AI’s transformative potential in optimizing traditional diagnostic tools, paving the way for more efficient and precise early ASD detection systems.

### 3.3. AI Techniques and Performance

The studies reviewed used various ML and DL techniques designed for specific types of data. Traditional ML models, like Naïve Bayes, Random Forest and Logistic Regression, worked well with structured datasets. Similarly, SVMs were highly effective in analyzing motion trajectories and neurophysiological signals, offering robust performance in tasks requiring precise feature extraction. For example, multi-kernel SVMs applied to dMRI data achieved notable accuracy, though they required extensive preprocessing and computational power. In contrast, DL models such as neural networks (like MNN, FNN, and ANN) performed better with unstructured and combined data types, delivering higher accuracy.

However, DL methods faced challenges like overfitting, high computational requirements, and the need for large datasets. To improve trust and usability, tools like SHapley Additive exPlanations (SHAP) and explainability algorithms were used by Paolucci et al., 2024 to make DL models more transparent, highlighting the balance between complex models and real-world clinical use.

### 3.4. Findings on Trade-offs and Multimodal Approaches

The studies highlight a trade-off between accuracy and practicality. Biomarker-based methods demonstrated high accuracy (e.g., AUC = 0.975) but required invasive procedures, whereas behavioral data approaches were more practical but less precise. Multimodal models, like those combining behavioral and neurophysiological data (Natraj et al., 2024), offer a promising direction by leveraging complementary strengths of different data types. However, these models face practical challenges, including the need for synchronized data collection and complex processing pipelines, which limit their scalability for clinical use.

## 4. Discussion

This review specifically focused on young children aged 0–7 years, a developmental period critical for identifying early markers of ASD. The rationale behind this age limit lies in the unique opportunity for early intervention during this time frame, which can significantly improve developmental outcomes. By restricting the scope to this age group, this review aimed to assess how AI has been applied to identify subtle early-life markers—such as gaze anomalies, motor irregularities, and neurophysiological patterns—and whether these methods effectively translate into practical early detection strategies.

The findings demonstrate that AI-driven tools, including portable biomarker devices and home-based video analysis systems, have shown potential in this age group, achieving high diagnostic accuracy (Hassanzadeh et al., 2023; Paolucci et al., 2023). AI tools are poised to revolutionize ASD Screening by improving efficiency and reducing reliance on subjective assessments. For example, integrating AI into established frameworks, such as the M-CHAT-R, has reduced diagnostic errors and streamlined workflows, offering tangible improvements over manual scoring (Achenie et al., 2019). Similarly, innovative modalities like tablet-based motion analysis and home video tools provide accessible, non-invasive diagnostic solutions. These approaches are particularly valuable in underserved regions, where traditional resources for ASD screening are often limited (Luongo et al., 2024; Paolucci et al., 2023).

AI technologies offer significant advancements in ASD screening as well, by improving precision, scalability, and accessibility. For instance, neural networks have achieved 94% accuracy in gaze and behavioral analysis, demonstrating AI’s ability to detect subtle markers often missed by traditional methods (Atyabi et al., 2023). Portable biomarker devices and home-based video analysis systems, as piloted in small-scale studies, hold promise for expanding diagnostic access to underserved regions (Jia et al., 2024). By automating complex analyses, these tools have the potential to democratize access to early ASD detection, ensuring timely interventions that can significantly improve developmental outcomes (Tonello et al., 2017).

However, real-world deployment is constrained by logistical and financial barriers, particularly in low-resource settings. Solutions such as cloud-based AI platforms can enable remote processing of biomarker or video data, reducing the need for on-site computational infrastructure. Partnerships with public health organizations and subsidized AI systems could further enhance access while mitigating resource constraints.

## 5. Limitations

Critical limitations identified include small sample sizes and demographic homogeneity, which increase the risk of overfitting and reduce generalizability. For instance, among studies analyzed cohorts of fewer than 50 participants, restricting their applicability to broader populations (Luongo et al., 2024).

Inconsistent validation methodologies also undermine the reliability of findings. Over-reliance on training datasets without sufficient external benchmarking compromises the robustness of AI models.

Despite the mentioned advancements, traditional methods retain advantages in terms of clinician trust and regulatory validation. AI tools often face skepticism due to their “black-box” nature, which obscures decision-making processes (Rudin, 2019). Explainability tools, such as SHAP values, have been developed to address this issue but are not yet widely adopted (Lundberg & Lee, 2017), but they remain unevenly and incompletely adopted in clinical workflows per recent healthcare XAI reviews (e.g., varied applications, limited critical evaluation/reporting). Bridging this gap will require ongoing collaboration between AI developers, clinicians, and policymakers to enhance the interpretability and reliability of AI models (Paolucci et al., 2023).

Furthermore, ethical and privacy concerns remain significant. Behavioral data (e.g., home videos used for algorithmic screening) can contain facial identities, household context, and voices of minors; neurophysiological data (EEG/eye-tracking) can be biometric and reveal sensitive traits. For example, ML classifiers on home videos have shown promising accuracy for ASD triage—raising stakes for secure storage, consent, and secondary use controls—while eye-tracking streams can reveal identity, age, mood, and even preferences; EEG has been studied for biometric authentication. These modalities therefore require stringent safeguards (access control, minimization, encryption, governance)

Beyond privacy, algorithmic bias is a central risk. Homogeneous training data (by sex, ethnicity, language, or site) can yield models that underperform for underrepresented groups (e.g., AFAB children and ethnic minorities), thereby exacerbating health inequities. Reviews of fairness in clinical ML document gaps in subgroup coverage, reporting, and evaluation; recent surveillance also shows persistent sex-linked prevalence differences that likely reflect measurement and access disparities rather than true incidence alone (Ueda et al., 2024).

Implementing AI-based diagnostic tools in healthcare often entails substantial costs, including expenses for advanced computational infrastructure, specialized equipment, and technical expertise. These financial demands can hinder the cost-effectiveness and scalability of such technologies, particularly in resource-limited settings. For instance, developing and deploying AI systems for healthcare can range from as low as $20,000 for a minimum viable product to upwards of $1 million for custom solutions (ScaleFocus, n.d.). Additionally, the requirement for specialized equipment and trained personnel further escalates costs, posing significant challenges for widespread adoption in low-resource environments (Zehra et al., 2023).

### 4.3. Future Directions

This review revealed that many AI-driven studies in this age range focus on detecting subtle markers, such as atypical gaze patterns or motor delays, which traditional methods may miss. However, gaps remain in ensuring the robustness and generalizability of these findings across diverse populations.

International collaborations should prioritize the development of large, inclusive datasets encompassing diverse ages, genders, and ethnicities to improve the generalizability of AI models (Jia et al., 2024). Such datasets can mitigate biases stemming from the demographic homogeneity observed in current studies, ensuring that AI tools equitably serve underrepresented populations, including females and ethnic minorities. Publicly accessible repositories can further facilitate cross-study comparisons and accelerate advancements in ASD research.

Integrating multiple data modalities, such as behavioral patterns, biomarkers, and neuroimaging, offers a promising pathway for enhancing diagnostic precision. By leveraging the strengths of each modality while mitigating individual limitations, multimodal systems can provide more comprehensive and reliable insights into ASD diagnostics (Natraj et al., 2024). Real-time synchronization protocols and cloud-based platforms can streamline the integration and analysis of diverse datasets, enabling seamless interoperability across diagnostic tools. To increase accessibility, especially in low-resource settings, scalable solutions of cloud-based platforms and smartphone-compatible tools should be prioritized. Portable systems, such as motion analysis games and biomarker testing kits, can bridge gaps in diagnostic availability by offering cost-effective and user-friendly options. These approaches can democratize access to early ASD diagnostics, particularly in underserved regions where traditional resources are scarce.

Future research should adopt appraisal frameworks such as PROBAST or CASP to ensure methodological rigor and consistency across studies. Standardized validation protocols can address inconsistencies observed in current AI models, particularly those relying heavily on training datasets without sufficient external benchmarking. By enhancing reliability, these frameworks can facilitate trust in AI-driven tools and promote their broader adoption in clinical practice.

Additionally, compliance with data privacy regulations is essential for safeguarding sensitive behavioral and neurophysiological data. Moreover, integrating fairness metrics into AI workflows, including bias audits and disparate impact analyses, can ensure equitable performance across diverse populations. Techniques such as federated learning and differential privacy can further balance the need for data sharing with robust security measures.

### 4.4. Conclusion

This review highlights AI’s potential to revolutionize early ASD diagnostics in preschooler children aged 0–7 years by identifying subtle markers like gaze anomalies and motor irregularities. AI tools excel in accuracy and efficiency across behavioral, biomarker, motion, and neurophysiological modalities, though challenges such as small sample sizes, demographic biases, cost-effectiveness and ethical concerns persist.

Future efforts must prioritize diverse datasets, scalable multimodal systems, and rigorous validation to ensure equity and reliability. With targeted advancements, AI can enhance diagnostic access and outcomes, transforming ASD screening and detection worldwide.

### 4.5. Funding

No specific funding was received for this work.

## Data Availability

This is a secondary Study. No data was made apart from PRISMA charts and Summary Table.

